# Emergence and Spread of SARS-CoV-2 Variants of Concern in Canada: a Retrospective Analysis from Clinical and Wastewater Data

**DOI:** 10.1101/2022.12.09.22283256

**Authors:** David Champredon, Devan Becker, Shelley W. Peterson, Edgard Mejia, Nikho Hizon, Andrea Schertzer, Mohamed Djebli, Yuwei Xie, Femi F. Oloye, Mohsen Asadi, Jenna Cantin, Markus Brinkmann, Kerry N. McPhedran, John P. Giesy, Chand Mangat

## Abstract

The spread of SARS-CoV-2 has been studied at unprecedented levels worldwide. In jurisdictions where molecular analysis was performed on large scales, the emergence and competition of numerous SARS-CoV-2 lineages has been observed in near real-time. Lineage identification, traditionally performed from clinical samples, can also be determined by sampling wastewater from sewersheds serving populations of interest. Of particular interest are variants of concern (VOCs), SARS-CoV-2 lineages that are associated with increased transmissibility and/or severity. Here, we consider clinical and wastewater data sources to retrospectively assess the emergence and spread of different VOCs in Canada. We show that, overall, wastewater-based VOC identification provides similar insights to the surveillance based on clinical samples. Based on clinical data, we observed a synchrony in VOC introduction as well as similar emergence speeds across most Canadian provinces despite the large geographical size of the country and differences in provincial public health measures. In particular, it took approximately four months for VOC Alpha and Delta to contribute to half of the incidence, whereas VOC Omicron achieved the same contribution in less than one month. By quantifying the timing and rapidity of SARS-CoV-2 VOCs invasion in Canada, this study provides important benchmarks to support preparedness for future VOCs, and to some extent, for future pandemics caused by other pathogens.

## 1 Introduction

Unprecedented clinical, genomic, and environmental surveillance applied to manage the on-going coronavirus disease 2019 (COVID-19) pandemic has allowed the scientific community to observe the emergence and spread of multiple lineages of severe acute respiratory syndrome coronavirus 2 (SARS-CoV-2). A consequence of the global prevalence of COVID-19 has been the emergence of SARS-CoV-2 variants, which have been closely monitored due to the acquisition of mutations that confer increased transmissibility or increased evasion of immune response functions [1, 2, 3, 4]. Variants associated with particularly high transmission and/or severity are designated “Variants of Concern” (VOC) by the World Health Organization and some have managed to propagate rapidly on a global scale. The first prominent and global VOC emerging in early 2021 was Alpha (lineage B.1.1.7), followed in June 2021 by Delta (lineage B.1.617.2), and since December 2021 Omicron (lineage B.1.1.529) has rapidly become the predominant strain. Some VOCs have not become dominant on a global scale, but have nevertheless reached a significant frequency at least regionally including Gamma in Brazil [5] and Beta in South Africa [2].

In addition to the traditional surveillance from clinical samples, wastewater-based epidemiology (WBE) can determine the proportion of VOC RNA in a given sewershed by use of sequencing or polymerase chain reaction (PCR) techniques [6, 7, 8]. Despite the inherent uncertainties in measuring viral concentration or estimating variants diversity in wastewater, WBE has been useful for informing public health officials and the general public on the status and trends of outbreaks [9, 10]. Hence, it is possible to use both data sources, clinical and wastewater, to potentially improve our confidence in estimates of VOC abundance circulating in a given population. Indeed, multiple international studies have shown that the relative abundance of a given VOC in wastewater correlates with the proportion of cases infected with this VOC identified by clinical surveillance [11, 12, 13, 14, 15, 16, 17, 18, 19, 20].

In light of the multiple SARS-CoV-2 lineages, a retrospective analysis of the dynamics of emergence and spread by VOCs – using wastewater and clinical data – can help inform preparedness planning for emergence of other VOCs in the future. Therefore, the objective of this study was to retrospectively review the invasion dynamics of the prevalent SARS-CoV-2 VOCs that have circulated in Canada, with the timing of emergence and speed of spread observed for those VOCs in several Canadian jurisdictions being quantified and described. Our analysis of VOCs circulation in Canada considers both clinical and wastewater data from VOC Alpha (the first to appear in Canada) up to the emergence of VOC Omicron (early 2022). Our results are in agreement with findings from studies from other countries.

As jurisdictions world-wide are transitioning to management of SARS-CoV-2 as an endemic virus, with accompanying reductions in health surveillance, a clear understanding on how an emerging VOC spreads at a regional and national scale will be useful for informing effective public health policy.

## 2 Methods

### 2.1 Clinical data

The Public Health Agency of Canada (PHAC) compiles a national line list of COVID cases that are reported by Canadian Provinces and Territories. For some records in this line list, the variant or lineage associated with each infection is identified. The lineage is either confirmed from whole genome sequencing or from “screening” via a discriminatory PCR assay that assesses the presence of variants by targeting specific mutations that are likely variant-defining, given the prior knowledge of circulating lineages nationally. In the event of both results being present, sequencing was prioritized over screening. These sequencing and screening efforts varied during the pandemic and across jurisdictions. In this study, this retrospective assessment focused on the four VOCs that reached non-negligible statuses in Canada: Alpha (lineage B.1.1.7), Gamma (lineage P.1), Delta (lineage B.1.617.2) and Omicron (lineage B.1.1.529). Thus, all other VOCs (*e*.*g*., Beta, Epsilon, Kappa) were ignored here. Because there could be some overlap with the mutations defining variants (*e*.*g*., Omicron and Alpha), we applied cut-off dates such that variants were assumed to exist only after those dates: 2020-11-01 for Alpha; 2021-02-01 for Gamma; 2021-03-01 for Delta; 2021-11-15 for Omicron.

### 2.2 Wastewater data

In October 2020, PHAC, the National Microbiology Laboratory (NML) and Statistics Canada started a pilot program that tested for the presence of SARS-CoV-2 in wastewater collected between two to three times per week from 15 wastewater treatment plants in various cities in Canada including Vancouver, Edmonton, Toronto, Montreal, and Halifax. In addition, wastewater was sampled from the three treatment plants in Winnipeg (the largest city in the province of Manitoba) five times a week through a provincial/municipal collaboration. All wastewater samples from Winnipeg, Vancouver, Edmonton, Toronto, Montreal and Halifax were shipped to NML in Winnipeg for qPCR analysis. To expand the population coverage of the NML/Statistic Canada program, we include the qPCR analysis of the wastewater sampled from Saskatoon (the largest city in the province of Saskatchewan) three times a week performed by the University of Saskatchewan (not NML).

Thus, the wastewater data samples the population of one large municipality in each province, whereas our clinical data samples the whole province. The comparison of the proportion of VOCs between clinical and wastewater data is still relevant because the catchment areas of the municipal wastewater treatment plants represent a substantial proportion of its provincial population. The population present in the catchment area of the wastewater treatment plants in Vancouver represents 48% of the total population of British Columbia; 24% of the province of Alberta for Edmonton; 23% of the province of Saskatchewan for Saskatoon; 54% of the province of Manitoba for Winnipeg; 20% of the province of Ontario for Toronto; 20% of the province of Quebec for Montreal; and 46% of the province of Nova Scotia for Halifax

In all sites, the proportion of the circulating VOCs present in a wastewater sample was quantified (*i*.*e*., not just a presence/absence test) by two laboratories only, the NML and the University of Saskatchewan (the latter analyzed the samples from Saskatoon only). Briefly, raw-post grit primary influent was shipped on cold-packs for testing to the NML (Winnipeg, Manitoba). Samples were stored at 4°C and tested within 1-3 days of receipt. Wastewater samples were mixed vigorously and a 30 mL aliquot was centrifuged (4000g, 4° C, 20 min) in a swinging bucket centrifuge. The pellet was then resuspended in Buffer RLT (Qiagen) containing 1% 2-mercaptoethanol and subjected to bead beating using a Bead Mill Homogenizer. The samples were then centrifuged (4000g, 4° C, 3 min) and total nucleic acids were extracted from the supernatant by use of the Roche MP96 instrument using the Magna Pure 96 DNA and Viral NA Large Volume Kit (Roche Diagnostics, Laval, QC) according to the Plasma External Lysis 4.0 protocol. See [21] for full methodological details. Base viral loads were determined by use of US-CDC N1/N2 assays [22]. The Alpha and Delta lineage assays were performed as previously described in [21]. The Omicron assay is described in Supplementary Material.

The raw influent samples from Saskatoon were processed as previously described in [17, 10] with modifications. Virus in 70 mL whole raw influent was enriched by PEG-8000 precipitation. Total RNA was extracted by use of RNeasy PowerMicrobiome Kit (Qiagen, Ontario, Canada). Percentage of Alpha VOC was determined by N D3L assay [23], while percentages of Delta and Omicron were determined by N200 assay [24]. There were no structured comparisons of laboratory performance between the NML (that analyzed all samples, except the ones from Saskatoon) and the laboratory at the University of Saskatchewan (that analyzed samples from Saskatoon only). However, the latter performed a comparison of concentration measurements between raw influent (the sample type used for Saskatoon in our study) and post grit primary influent samples (the sample type used by NML in our study), all collected in Saskatoon, and found a significant correlation (*R*^2^ = 0.57, *p <* 0.001, manuscript in preparation by Oloye *et al*.). Moreover, the respective methods of each laboratory remained the same during the study period which allows us to perform the longitudinal analysis presented here, despite potential differences between the two laboratory assays.

### 2.3 Statistical analysis

Using the PHAC line list, clinical cases that had been identified as one of the four VOCs of interest (through screening or sequencing) were retained. Frequencies of circulation of variant *v* at time *t* is therefore defined as *p*(*v, t*) = *n*_*v*_(*t*)*/n*(*t*) where *n*_*v*_(*t*) is the number of clinical cases identified with variant *v* on day *t, n*(*t*) is the number of clinical cases where any variant was identified on day *t*. A multinomial distribution for *n*_*v*_ was assumed. Confidence intervals for proportions of each VOC were simultaneously estimated by use of the function `MultinomCI` from the R [25] package `DescTools` version 0.99.44 [26]. To evaluate growth rates (*i*.*e*., speed of initial spread) of *clinical* data during each wave of a given VOC in a given jurisdiction, we assumed the time-dependent proportions of VOCs followed a logistic growth model. The three parameters defining the logistic function – location, steepness and asymptotic value – were estimated from clinical data only. We fit the models independently for each province (using only clinical data) with Markov chain Monte-Carlo implemented in the R library `rjags` version 4-12 [27]. To calculate the time since introduction in the logistic growth model, we assumed a single date of introduction nationwide for each VOC : 2020-11-01 for Alpha, 2021-02-01 for Gamma, 2021-03-01 for Delta and 2021-11-15 for Omicron. Moreover, to aggregate provincial clinical data into national estimates, a similar logistical growth model was applied, but with a hierarchical structure. A full description of the statistical models is given in the Supplementary Materials.

The VOC proportions estimated from wastewater are simply the variant allele percentages detected by the corresponding assay. No confidence intervals were derived. Moreover, wastewater data was not used to fit the logistic models due to insufficient data at the beginnings of most waves of SARS-CoV-2 associated with various VOCs. Hence, growth rate of each VOC will be assessed using clinical data only. However, the wastewater-based VOC proportions will be compared to the clinical-based ones to assess their similarity.

## 3 Results

Findings are reported for seven Canadian provinces that had a sufficiently large sample size of VOC identified to provide interpretable results: British Columbia (B.C.), Alberta (Alta.), Saskatchewan (Sask.), Manitoba (Man.), Ontario (Ont.), Quebec (Que.) and, Newfoundland and Labrador (N.L.).

The trends of VOC proportions observed from the clinical data (shaded areas, Figure 1) and from the wastewater data (thick lines, Figure 1) appear similar in all provinces. However, some regional differences were observed. VOC Gamma occurred at a significant frequency only in British Columbia and Saskatchewan, but did not displace the emergence of VOC Delta (Figure 1). During summer 2021, three VOCs, Alpha, Delta and Gamma, probably co-circulated at substantial levels in Alberta, British Columbia and Saskatchewan, whereas transitions between these VOCs appeared to be faster in Ontario and Quebec. Differences in sizes of clinical samples (Figure S1) affected the size of uncertainties in proportion estimates and result in confidence intervals of different sizes (*i*.*e*., confidence intervals broadening when the sample size is small. Figure 1, shaded areas). The proportions of all clinical samples reported that could not be assigned to any VOC, due to screening results being unable to distinguish between circulating VOCs or sequencing results not being classified as a VOC, was variable in time and across provinces (Figure S2). The proportion of unidentified VOCs in the clinical samples was generally below approximately 80% in each province, except during the early phase of the VOC Alpha and after July 2021 in Quebec and Manitoba (Figure S2). Hence, we generally had a sufficient sample size to estimate VOC proportions from clinical samples (the uncertainty being reflected in the width of the confidence intervals in Figure 1).

**Figure 1:**
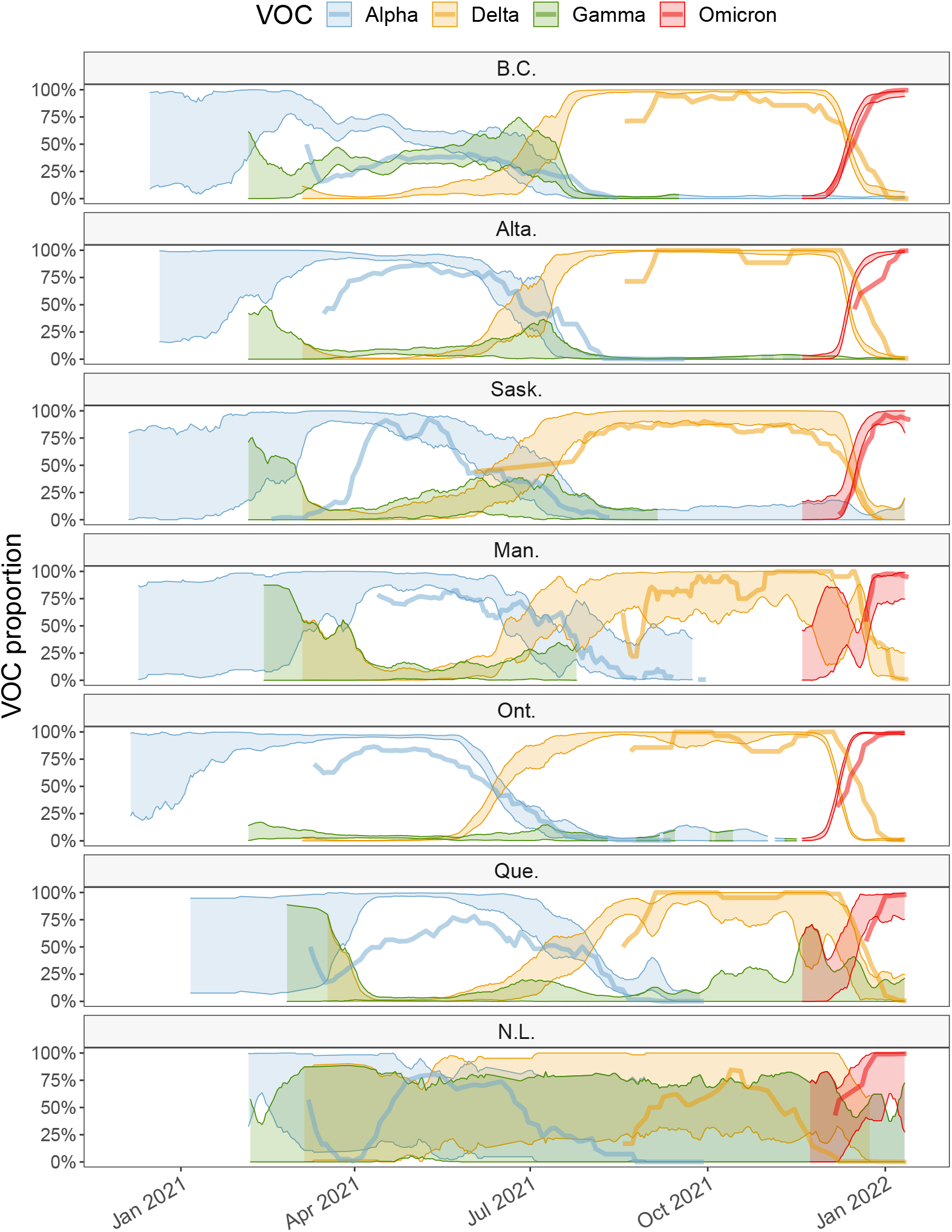
Proportions of VOCs circulating in each province, estimated from the sequencing of clinical samples (shaded area representing the upper and lower 95% confidence interval) and RT-qPCR applied to VOC-defining mutation targets from wastewater samples (thick solid line, representing the mean estimate). Data sources: PHAC (clinical surveillance); PHAC/NML (wastewater surveillance for B.C., Alta., Man., Ont., Que. and N.L); University of Saskatchewan (wastewater surveillance for Sask.). Delta assay was only available after 18-Aug-2021. Abbreviations: British Columbia (B.C.), Alberta (Alta.), Saskatchewan (Sask.), Manitoba (Man.), Ontario (Ont.), Quebec (Que.) and, Newfoundland and Labrador (N.L.)

In most provinces, proportions of VOCs measured from wastewater samples (Figure 1, thick lines) show similar trends as the proportions estimated from clinical surveillance, especially for the Omicron VOC. The assay applied for Delta in wastewater by NML was available only after 2021-08-18, hence there is no wastewater-based estimates of Delta proportions before this date (except for Saskatoon). Proportions of VOCs observed in municipal wastewater only are shown in Figure S3.

The fit of the logistic growth model on *clinical* data of VOCs observed during their initial introduction phase is shown in Figure 2. Credible predictive intervals for VOC Alpha are broad because clinical data were collected too late to capture the early invasion stage (Figure 2). According to the logistic fit for VOC Gamma we infer that this VOC circulated at significant levels only British Columbia.

**Figure 2:**
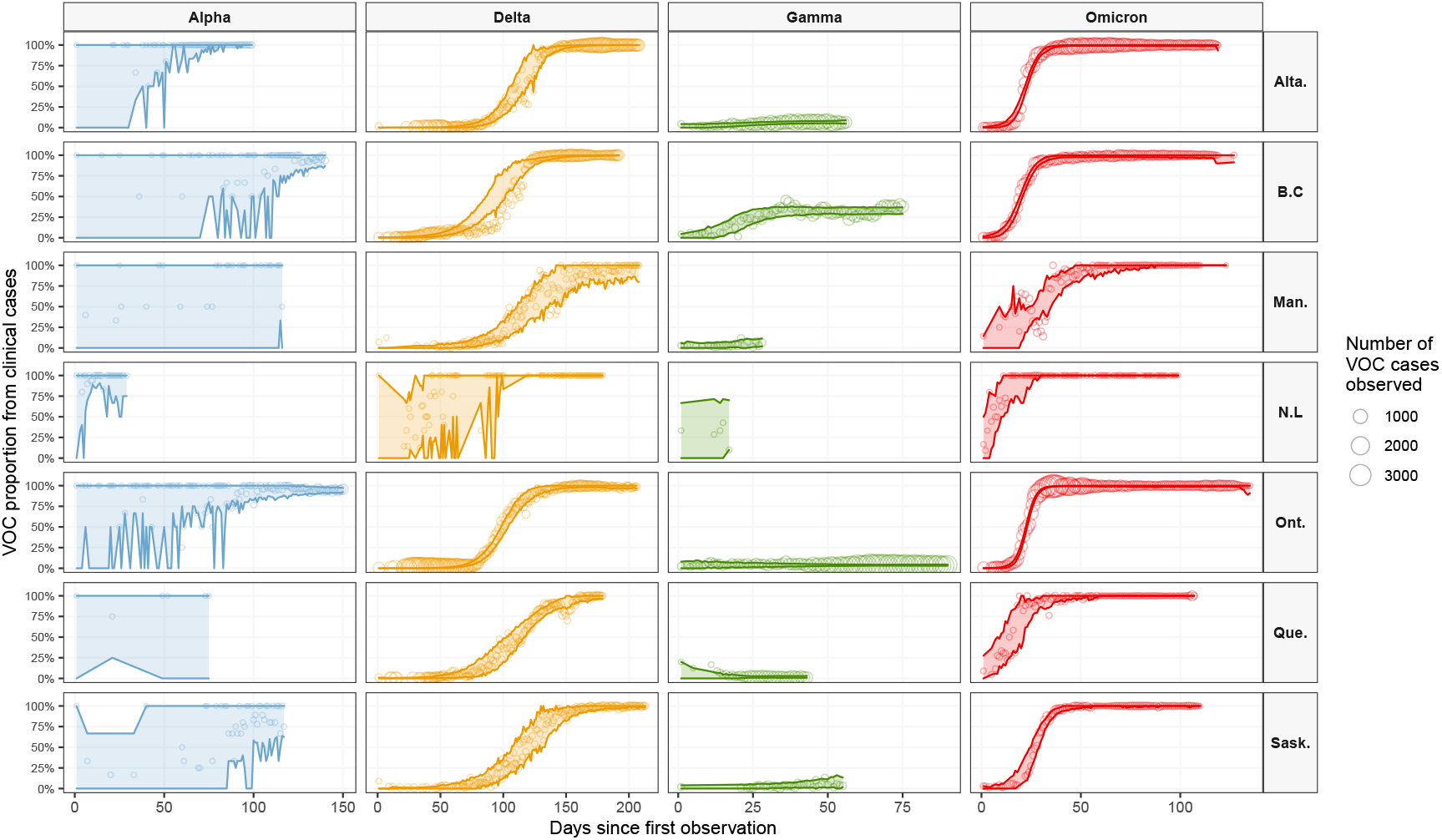
Logistic growth model fit to initial invasion. The circles represent the observed proportion of the VOC; circle size is proportional to the number of clinical samples identified as that VOC. The shaded area and solid lines represent the upper and lower bounds of the 95% credible intervals for prediction of the logistic growth model. Abbreviations: British Columbia (B.C), Alberta (Alta.), Saskatchewan (Sask.), Manitoba (Man.), Ontario (Ont.), Quebec (Que.) and, Newfoundland and Labrador (N.L)

Alpha, Delta and Omicron were the only VOCs that successfully invaded all jurisdiction. The uncertainty for VOC Alpha is very large because we have enough data one the initial introduction phase for Delta and Omicron only (Figure S2). Speeds of spread for these two VOCs were similar among provinces as estimated as time after the first observation in Canada to reach a proportion of 50% (Figure 3). The estimate for VOC Delta in Newfoundland is an outlier and probably affected by the small sample size.

**Figure 3:**
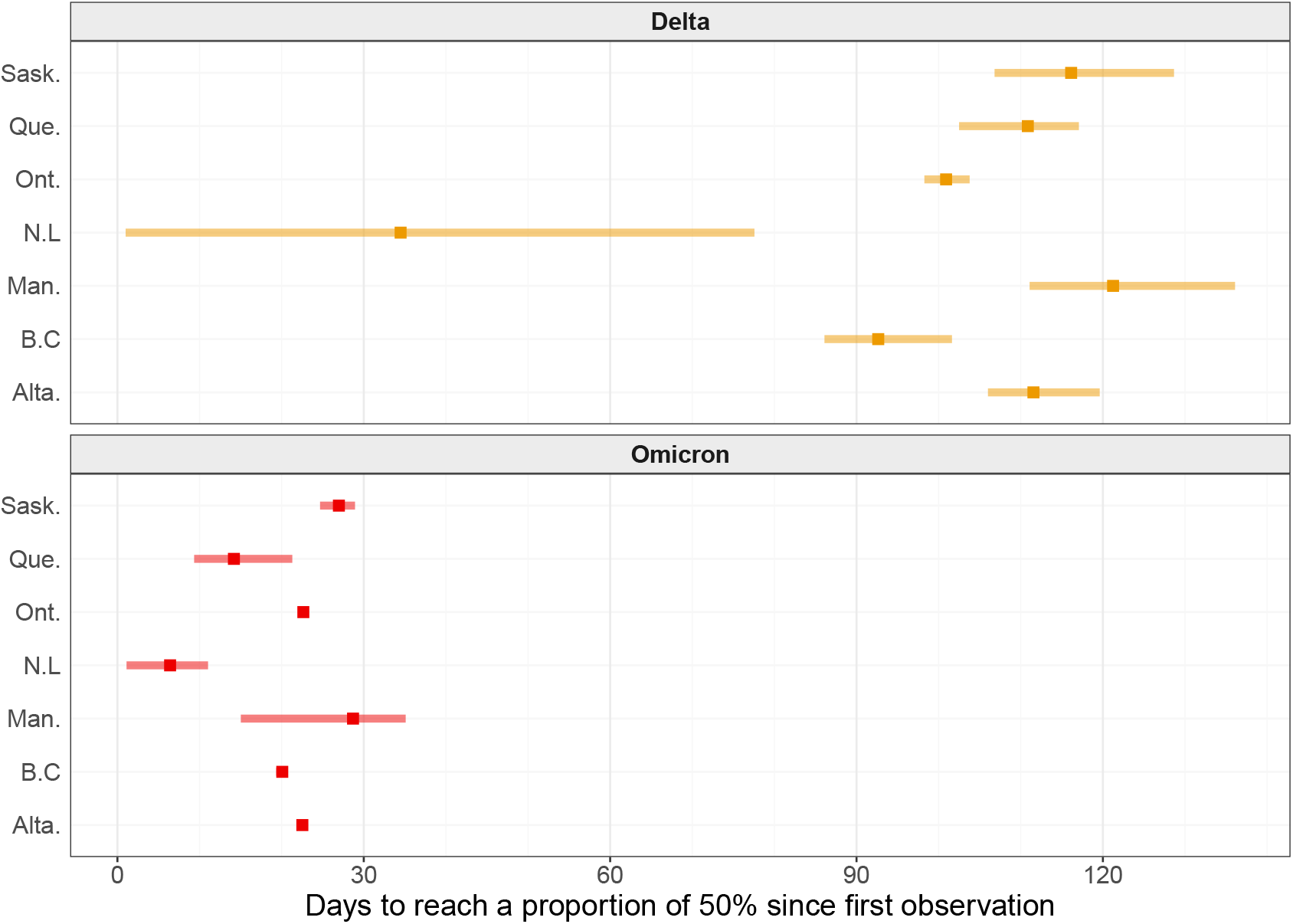
Time taken for each VOC to reach a proportion of 50% after its first observation in the respective province. Point represents the mean estimate and the Width of the horizontal blue segment is the predictive 95% credible interval.

VOC Delta took between three and four months to become the major circulating variant in Canada, but VOC Omicron required less than one month to become dominant. The same results displayed with respect to calendar dates are presented in Figure S4.

The fit of the hierarchical model to clinical data provides a national perspective of the initial invasion dynamics for each VOC. Posterior estimates presented in Figure 4 suggest that Alpha had nearly fully invaded Canada by early January 2021, Delta by early July 2021, and Omicron by early January 2022. Despite its inability to invade fully, Gamma VOC probably reached a maximum proportion of about 25% nationally.

**Figure 4:**
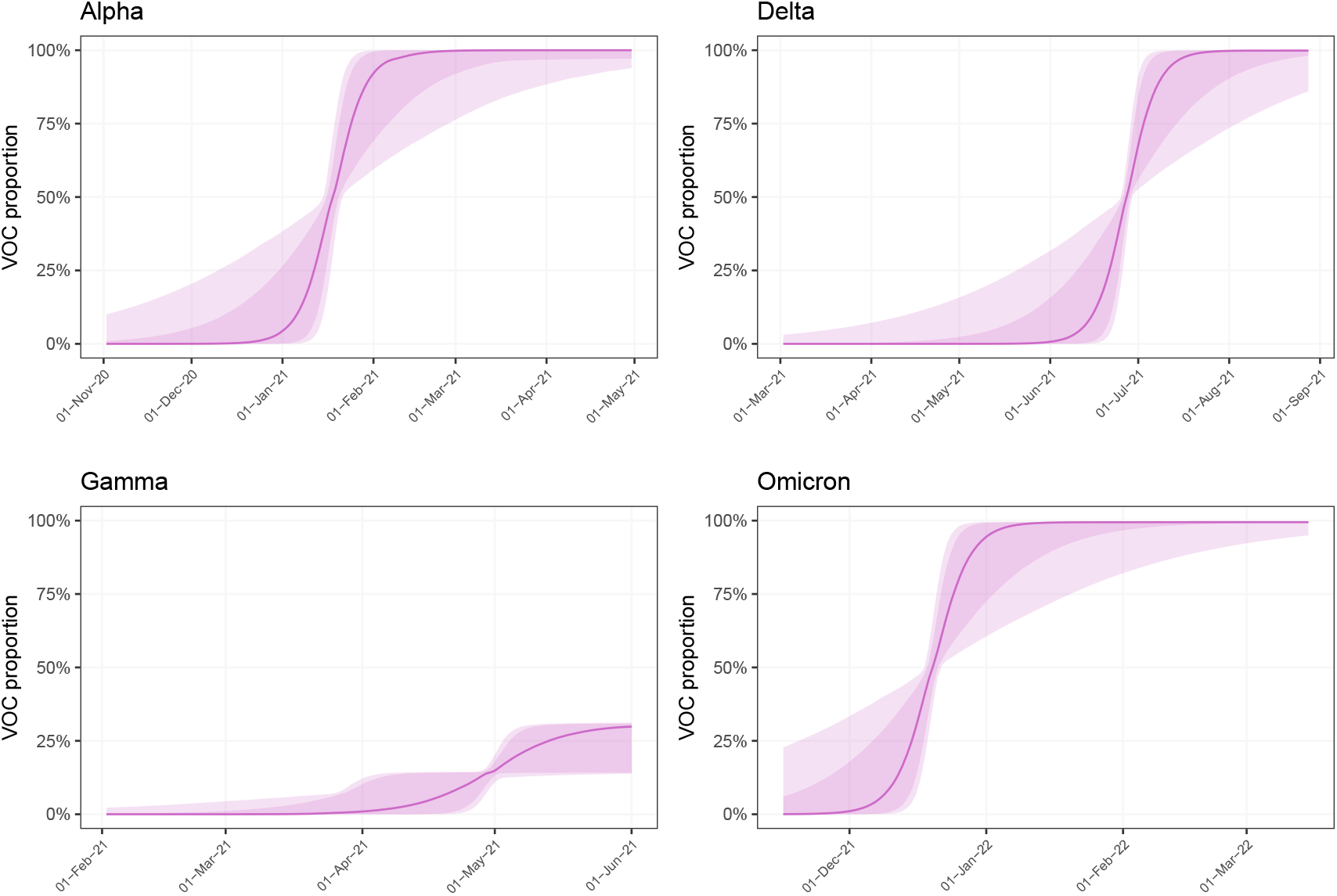
Estimates of circulating VOC proportions in Canada. Thick line represents the mean, dark (resp. light) shaded area the 80% (resp. 95%) credible interval.

## 4 Discussion

The COVID-19 pandemic has provided the first opportunity to observe, in detail and near real time, an emerging pathogen spreading in human populations globally. Moreover, due to unprecedented sequencing efforts with over 8 million clinical sequences collected worldwide and uploaded to the sharing platform GISAID (www.gisaid.org) in less than two years, it has been possible to track the evolution of SARS-CoV-2. This includes observing emergences of various lineages and describing how they have spread in various populations, in particular their rate of spread and severity of illness.

This study reports the spread of four SARS-CoV-2 variants of concern (VOCs) across seven Canadian provinces by use of clinical and wastewater surveillance data, covering approximately 95% and 25% of the population of Canada, respectively. Despite being a geographically large country, in which public health measures related to COVID-19 were decided and implemented at the provincial level, synchrony in the VOC introduction was observed in all jurisdictions with similar speeds of invasion for each VOC. This is remarkable since there were notable differences in management of COVID-19 by implementation of various public health measures between provinces [28]. Despite having a small sample size (due to its population size) Newfoundland and Labrador appeared to be an outlier. This province had very different public health measures, at least as it pertained to the movement of people, and it managed to keep the number of COVID-19 cases relatively low, at least until the Omicron wave. At the time when Delta started to invade, there were virtually no local transmission in this province. Overall, although public health measures probably helped to limit locally COVID-19 morbidity and mortality, differences in those measures among jurisdictions did not appear to have large effects on the emergence or the speed of spread of VOCs. This might have been caused by simultaneous importation events (*e*.*g*., international travel linked to essential businesses and services never stopped in Canada) and the intrinsic high contagiousness of SARS-CoV-2 even among asymptomatic cases. Large uncertainties in estimates of proportions of VOCs, which result in broad confidence intervals, underscore the importance of having sufficiently large and robust programs to surveil lineages to accurately monitor VOC emergence and spread. The proportion of clinical samples without VOC identification was variable in our dataset (Figure S2) and not explicitly modelled, hence periods with a low proportion should broaden even more our confidence intervals. Moreover, such a surveillance should rely, partially, on random sampling to avoid biased estimates.

Effective reproduction numbers (also known as “*R*_*t*_”), which are critical metrics to inform public health actions but lack the time dimension, have been estimated for VOCs in various Canadian locations [29]. It is possible to estimate *R*_*t*_ using both clinical and wastewater data [30, 31]. Here, estimates based on a logistic growth model complement *R*_*t*_ metrics (not estimated in this study) and provide practical, easy-to-understand, quantitative information about the speed of initial spread. Rates of spread reported here provide benchmarks for what to expect from future SARS-CoV-2 VOCs that might occur.

VOCs Delta and Gamma, both having few novel mutations, took about four months to become predominant. However, Omicron with multiple novel mutations on the spike protein reached a proportion of 50% in less than a month. Our findings are similar to those of other studies have found in Canada [32, 33, 34] and in other countries [11, 12, 13, 16, 18, 20]

These are important metrics that can assist in anticipating the necessary public health reaction time should other VOCs emerge in the future. This study uses two data streams: clinical reports of COVID-19 cases (used to fit statistical models) and SARS-CoV-2 concentration in wastewater (for comparison). Since they each have different sources of bias, the approximate agreement between both sources strengthens inferences of dynamics of relative proportions of VOCs in Canada. Clinical sampling is usually not random, including in Canada. Some socio-demographic groups, such as healthcare professionals, might be over-represented. Outbreaks may also be over-represented. Other groups might have limited access to testing and sequencing capacity might be limited during large infection waves, which might exacerbate these and other biases. Clinical testing strategies (including capacity, and targeted populations) also varied by jurisdiction. Wastewater sampling might be affected by environmental events, such as large rainfalls or snowmelts, industrial wastes, and events causing a large influx of people into a sewershed, such as sporting events or conferences. Finally, areas studied were limited to major urban centers and did not reflect the burden of disease in remote regions of Canada, except for Newfoundland and Labrador.

The agreement between clinical and wastewater data, especially that observed during the Omicron wave, validates the ability to monitor proportions of VOCs circulating in communities from data collected in wastewater [35, 10, 36]. The WBE estimates can also provide an opportunity to better triangulate VOC proportions in locations where fewer clinical samples have been available, such as Newfoundland and Labrador (Figure 1, where larger uncertainty from clinical data is indicated by the widths of confidence intervals). Even if it does not reach the level of precision of clinical surveillance, WBE based on PCR assays targeted on VOC-defining mutations has the potential to be a cost-effective surveillance method that may be more consistent both in time and across jurisdictions (especially when the wastewater analysis is performed using a stable assay in each laboratory in order to perform a longitudinal analysis – as it is the case here in this study). Although currently not timely enough to provide a near real-time surveillance, full sequencing of SARS-CoV-2 genetic materials found in wastewater can complement PCR-based assays because it provides a more complete and detailed picture of circulating VOCs, and can potentially identify new dominant variants (but the development and validation time for new wastewater-based assays may hamper timeliness). There is a clear advantage to perform full sequencing (vs. PCR-based assay) when there are multiple overlapping mutations defining different lineages, as this is the case in the post-Omicron era.

Interpretation of differences between proportions of various VOCs estimated from clinical and wastewater data is not evident. Indeed, a clinical surveillance that indicates a larger proportion of a particular VOC than that determined in WBE estimation, for example Alpha during spring 2021 in Ontario and Quebec, could be caused by a bias where clinical samples were preferentially selected among patients infected with Alpha because they were more symptomatic compared to wild types. Other factors that could bias VOC proportions can be periods of enhanced testing for contacts by some jurisdictions and preferential sequencing for travelers from certain regions. Alternatively, the lesser proportion estimated from WBE might be caused by lesser sensitivity of the laboratory assay. There is also an intrinsic demographic mismatch between clinical surveillance, which is performed province-wide, and wastewater samples that may only be collected in only a few large cities in each province (in this study, the single largest city). Hence, differences between proportions of VOCs estimated from clinical and wastewater data can also be caused by a VOC emergence that might not occur at the same time and speed in the largest cities of a province as it does in the whole province. Finally, the small sample size of clinical samples experienced at times in some provinces (Figures S1 and S2) may have impacted the accuracy of our analyses.

In conclusion, based on clinical surveillance data the SARS-CoV-2 VOCs have synchronously invaded different parts of Canada despite being distant and with various intensities of independently implemented COVID-19 public health measures. Wastewater-based surveillance can be a good indicator of VOCs circulation in communities and can complement clinical surveillance. Comparative estimates of the rapidity of spread and synchrony for past waves driven by VOCs can support preparedness for next VOCs and, to some extent, next pandemics from other pathogens.

## Supporting information

Supplement - Statistical model

## Data Availability

All data produced in the present study are available upon reasonable request to the authors

## Acknowledgments

We thank the provincial public health agencies that collected and shared the clinical data.

We thank Statistics Canada’s Canadian Wastewater Survey team and municipal partners for i) providing wastewater samples, and ii) processing and compiling wastewater metadata.

The data from Saskatchewan was collected as part of the project titled “Next generation solutions to ensure healthy water resources for future generations” funded by the Global Water Futures program, Canada First Research Excellence Fund (#419205; Additional information is available at www.globalwaterfutures.ca) and the Public Health Agency of Canada. Dr. Brinkmann is currently a faculty member of the Global Water Futures program. The authors acknowledge support of the City of Saskatoon Wastewater Treatment Plant. The research was supported by Discovery Grants from the Natural Science and Engineering Research Council of Canada awarded to Prof. Giesy and Dr. Brinkmann. The authors wish to acknowledge the support of an instrumentation grant from the Canada Foundation for Infrastructure. Prof. Giesy was supported by the Canada Research Chairs Program of the Natural Sciences and Engineering Research Council of Canada (NSERC) and a distinguished visiting professorship of Environmental Sciences from Baylor University in Waco, Texas, USA.

We acknowledge Dr. Anil Nichani and Aamir Fazil for facilitating the wastewater surveillance and wastewater-based modelling activities at PHAC/NML.

## Contributions of Authors

DC: wrote initial manuscript, design study, performed part of statistical analysis

DB: performed Bayesian statistical analysis

SP, EM, NH: data collection, methodology, validation, data analysis

CM: project management, method development, data collection, provided funding, scientific oversight

JPG, MB, KNM: provided funding, project management, method development; data collection and curation; editing of final manuscript

YX, FFO, MA: method development, data collection and curation, editing of final manuscript

JC: Data collection and curation

