## Supplementary figures and images for "Emergence and Spread of SARS-CoV-2 Variants of Concern in Canada: a Retrospective Analysis from Clinical and Wastewater Data"

### Figure S1

Figure S1 − Number of clinical samples sequenced or screened, by jurisdiction

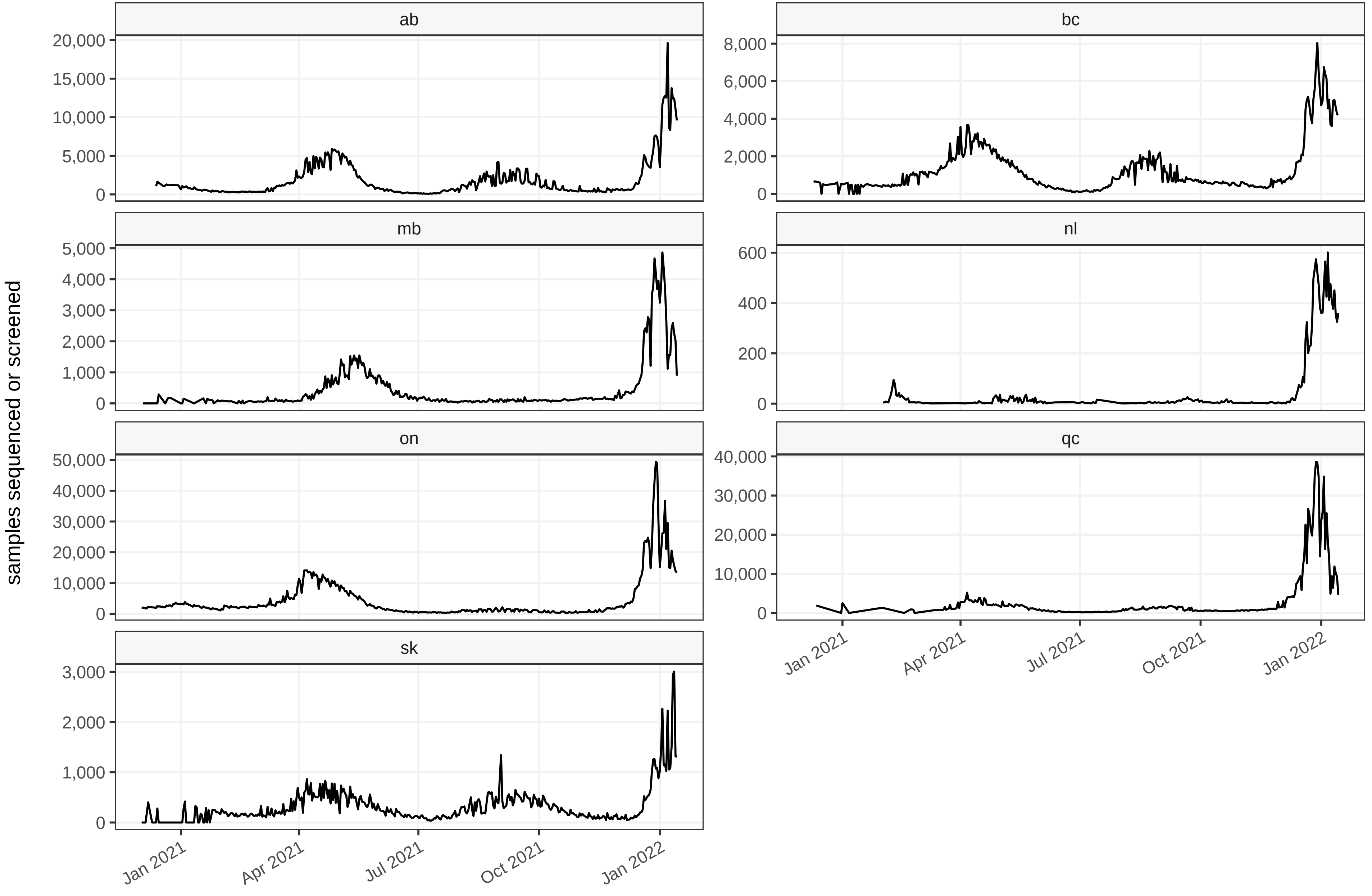

### Figure S2

Figure S2 − Proportion of all clinical samples without VOC identification

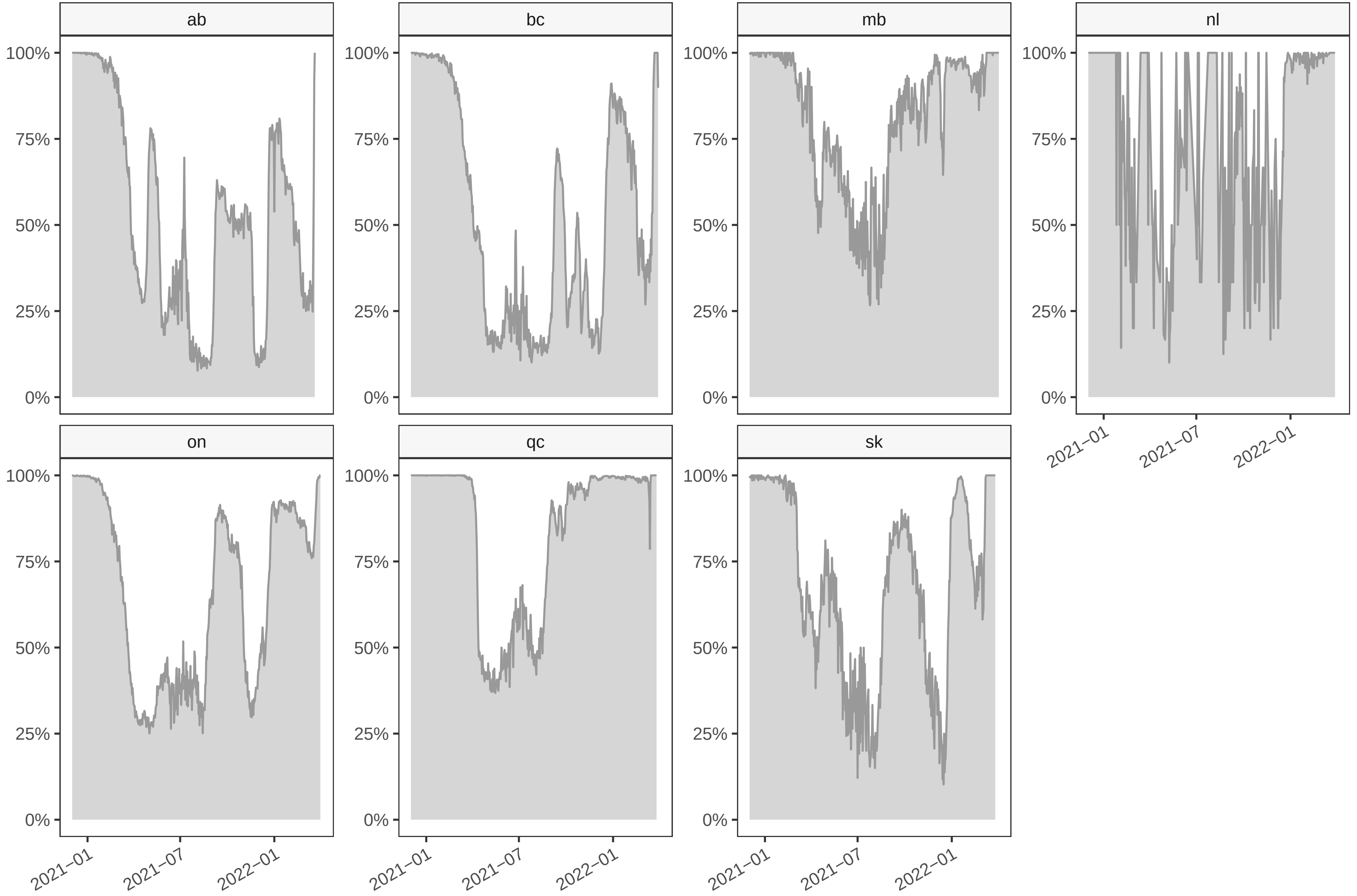

### Figure S3

#### Figure S3 − Proportion of VOC from wastewater samples only

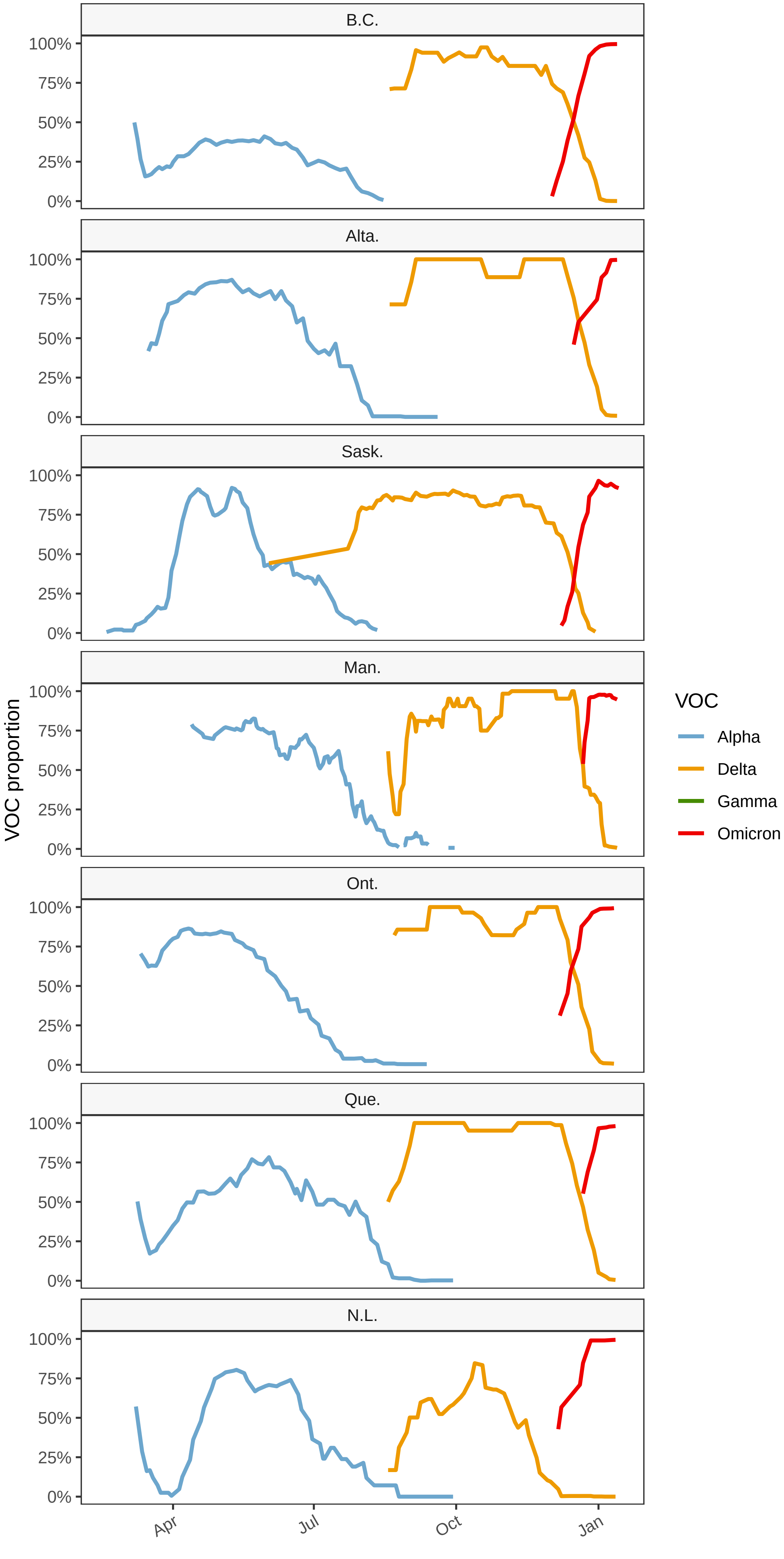

### Figure S4

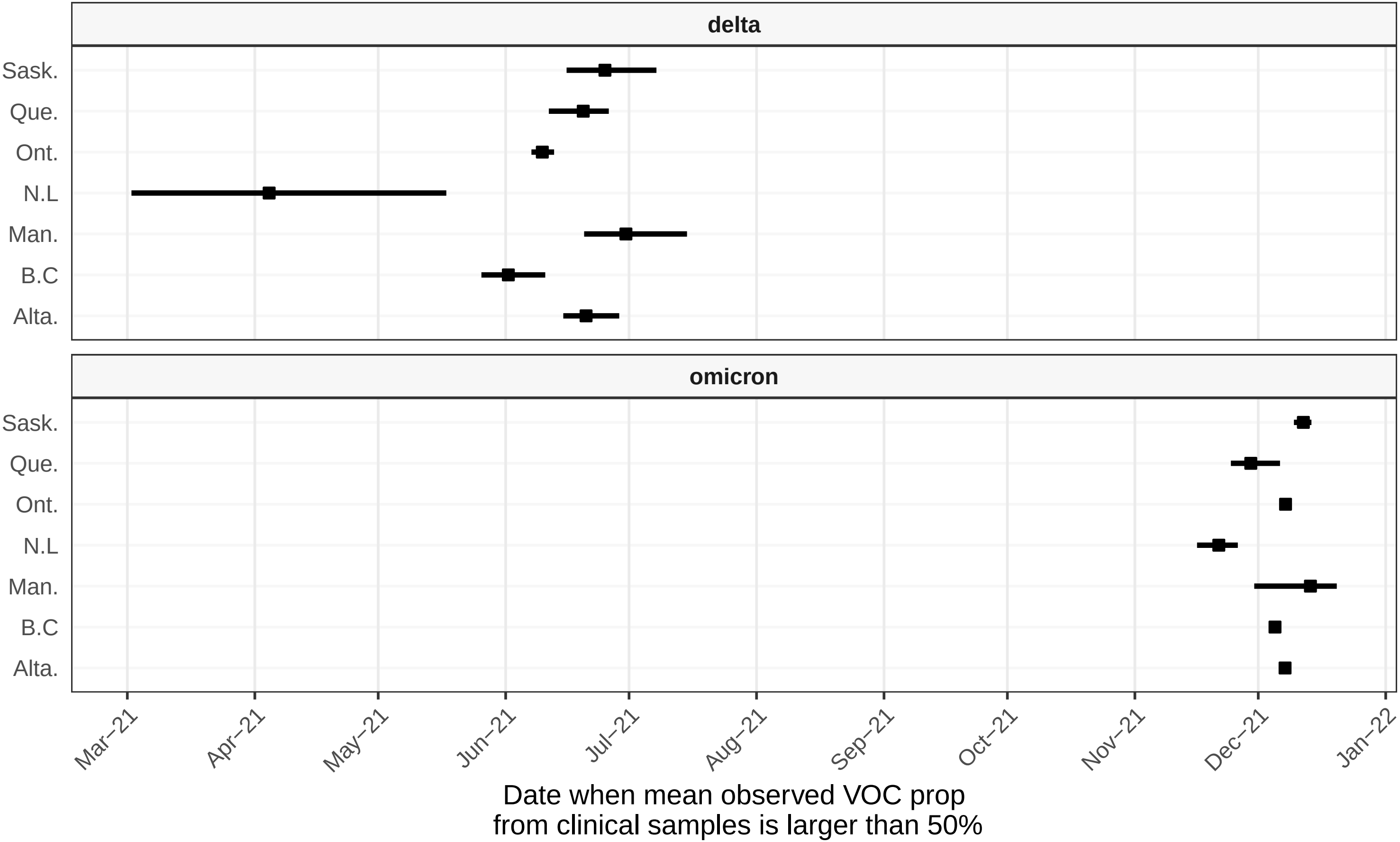
