## Supplement - Statistical model for "Emergence and Spread of SARS-CoV-2 Variants of Concern in Canada: a Retrospective Analysis from Clinical and Wastewater Data"

### Logistic Growth Model for VOC Proportions

Devan Becker, David Champredon

We used two logistic growth models for this analysis: One fit to each province/variant combination separately and one that “shares strength'' across provinces. Both models assume that, at time $t$ (measured as an integer number of days since the first appearance in the data), the proportion of cases that are caused by variant $\nu$ in province $j$ is $p_{j\nu}(t)$. Both models also assume that $p_{j\nu}(t)$ can be modelled using a logistic growth curve:

$$p_{j\nu}(t) = \frac{K_{j\nu}}{1 + \exp((m_{j\nu} - t)r_{j\nu})} \tag{1}$$

The parameter interpretations are as follows. $K_{j\nu}$ is the asymptote (the value of the growth curve as $t$ approaches infinity) for the proportion in province $j$ for variant $\nu$. $m_{j\nu}$ is the midpoint of the curve, which is the time point at which 50% of the cases in province $j$ are attributed to variant $\nu$ (it is also the time at which the curve reaches it's maximum slope). Finally, $r_{j\nu}$ is the rate parameter, which controls how steep the curve is.

We fit these data with a binomial likelihood where the number of cases of variant $\nu$ is labelled $Y_{j\nu}(t)$ and the total number of cases with any known variant is labelled $N_{j\nu}(t)$. This formulation of the likelihood incorporates the correct uncertainty in the model - when $N_{j\nu}$ is small, the uncertainty is higher (and therefore the credible intervals are larger). This model is fit in a Bayesian setting, with the following likelihood and prior distributions:

$$\begin{aligned}
Y_{j\nu}(t) &\sim \text{Binomial}(p_{j\nu}(t), N_{j\nu}(t)) && (2)\\
p_{j\nu}(t) &= \frac{K_{j\nu}}{1 + \exp((m_{j\nu} - t)r_{j\nu})} && (3)\\
K_{0j\nu} &\sim \text{Bernoulli}(0.7) && (4)\\
K_{1j\nu} &\sim \text{Beta}(6, 2) && (5)
\end{aligned}$$

$$
\begin{aligned}
K_{j\nu} &= K_{0j\nu}K_{1j\nu} + (1 - K_{0j\nu}) && (6)\\
m_{j\nu} &\sim \text{Normal}(m^*_{j\nu}, 5^2)T(0,) && (7)\\
r_{j\nu} &\sim \text{Gamma}(2, 2) && (8)
\end{aligned}
$$

In the model specification above, note that a variation on a spike-and-slab prior [1, 2] is used for the asymptote so that the posterior may have point mass at 1 (i.e. it is possible that all posterior draws are exactly 1). In the specification of the midpoint prior, note that $T(0,)$ indicates truncation with a lower bound at 0 with no specified upper bound.

Our two models differ in the specification of $m^*_{j\nu}$: when the provinces are modelled separately, this is specified as the mean of the observated dates for a given province/variant; to share strength between provinces, $m^*_{j\nu}$ is modelled with a normal prior distribution, again centered at the mean of the observed dates for a province and with a variance of $5^2$ and constrained to be positive. By modelling it the second way, the midpoint for each province is modelled as a random deviation from a country-wide effect. Because of imbalanced sample sizes across provinces, the posterior distributions for the midpoint in larger provinces are dominated by the data while smaller provinces borrow more information from the country effect.

The models were run using the JAGS [3] software within the R software environment [4]. We ran 3 chains, each with 5,000 initial iterations that were not used (burn-in iterations) and then used every 10th value of the next 10,000 iterations ("thining"). Convergence was confirmed by the upper bounds of the Gelman-Rubin statistics as well as visual inspection of the trace plots.

[4] R Core Team. R: A language and environment for statistical computing. 2021.
